# Mass Screening for SARS-CoV-2 Infection among Residents and Staff in Twenty-eight Long-term Care Facilities in Fulton County, Georgia

**DOI:** 10.1101/2020.07.01.20144162

**Authors:** Carson Telford, Udodirim Onwubiko, David Holland, Kim Turner, Juliana Prieto, Sasha Smith, Jane Yoon, Wecheeta Brown, Allison Chamberlain, Neel Gandhi, Shamimul Khan, Steve Williams, Fazle Khan, Sarita Shah

**Author notes:** **Contact information:** Carson Telford, Fulton County Board of Health, 10 Park Place South SE, Suite 427, Atlanta, GA 30303.

## Abstract

Mass screening for SARS-CoV-2 infection in long-term care facilities revealed significantly higher prevalence of infection in facilities that screened in response to a known infection compared to those that screened as a prevention measure. “Response” facilities had a SARS-CoV-2 prevalence of 28.9% while “preventive” facilities’ prevalence was 1.6% (p <0.001).

## Background

Since the first reported case of Coronavirus Disease 2019 (COVID-19) in the United States in January 2020, there have been more than 2.3 million cases and 120,000 deaths as of June 22, 2020. The disease is associated with substantially higher mortality among the elderly and those who have comorbid medical conditions; the risk of death is 10-20 times higher among persons over age 70 compared to those under age 50 across diverse epidemiologic settings [1]. Long-term care facility (LTCF) residents are at particularly high risk for SARS-CoV-2 infection and death given their advanced age and high prevalence of chronic medical conditions, combined with functional impairment that necessitates frequent, close contact with healthcare providers, who may inadvertently spread disease among residents. Despite comprising <1% of the U.S. population, a staggering 42% of COVID-related deaths are among LTCF residents [2]. Several outbreaks among LTCFs have been reported, most notably the earliest description from Washington state of 167 cases linked to a single facility, with a case fatality of 33.7% among residents [3].

Mass screening for SARS-CoV-2 infection in high-risk populations has been proposed as an efficient method for identifying those most likely to have severe disease and for curtailing widespread transmission among vulnerable individuals [4]. Because nearly half of persons with COVID-19 can be asymptomatic or presymptomatic, but still infectious, active screening can facilitate early identification, supportive treatment, and enhanced infection control measures [5]. To date, more than 450 LTCFs in Georgia have reported at least one COVID-19 case; LTCF residents comprise 10% of all cases reported (N=6,591), but account for 45% of all deaths with a COVID-19 diagnosis [6]. Thus, evaluating the outcomes of active screening initiatives in LTCFs is a high priority.

In conjunction with the Georgia Emergency Management Agency and the National Guard, the Fulton County Board of Health (FCBOH) has conducted mass screening in LTCFs since March 2020. We report results from 15 LTCFs where mass screening was conducted among residents and staff after cases had been reported and 13 LTCFs where preventive mass screening was conducted (i.e., where no cases had yet been reported).

## Methods

Fulton County includes the city of Atlanta, with a total population of approximately 1.1 million persons in 2019. Mass screenings of residents and staff in LTCFs began in March 2020 and initially occurred in response to reports of LTCF residents who were hospitalized with COVID-19 symptoms and thereafter tested positive for SARS-CoV-2 virus (i.e., “Response group”). Due in part to increasing outbreaks identified in LTCFs across Georgia, supplemental support from the National Guard was added in April 2020 to enable mass screening of LTCFs irrespective of prior COVID-19 case identification. LTCFs which underwent mass COVID-19 screenings of staff and residents as a preventive measure (i.e. prior to identification of an active case) are considered the “Preventive group”.

Nasopharyngeal swab samples were collected by trained healthcare staff from FCBOH and transported to various Georgia laboratories to be tested using Reverse Transcription Polymerase Chain Reaction (RT-PCR). Six facilities contracted private companies to collect nasopharyngeal swabs and test samples for residents and staff using RT-PCR, then reported results to the FCBOH. Collection, transportation, and testing of samples were conducted in accordance with the most recent CDC guidelines [7]. All results were reported back to the facilities, where residents and staff were referred for appropriate care depending on test results [8].

We defined LTCF as any facility which provides skilled nursing services, including nursing homes, memory care, and assisted living facilities located in Fulton County. A confirmed case was defined as a person with a positive SARS-CoV-2 RT-PCR result. We included all confirmed COVID-19 positive individuals in our final case count per facility, regardless of whether they were diagnosed through mass screening or through other testing (e.g., as part of diagnostic evaluation of respiratory illness).

Frequency distributions (counts and percentages) were used to describe COVID-19 positivity, hospitalizations, and deaths among residents and staff. Fisher’s exact test was used to test differences between facility groups and p<0.05 was used as the cut-off for statistical significance.

This activity was reviewed by the Georgia Department of Public Health and deemed exempt from IRB review as a public health surveillance activity in response to the COVID-19 emergency response.

## Results

Mass testing was conducted in 28 LTCFs where a total of 5,671 individuals (2,868 [50.6%] residents and 2,803 [49.4%] staff) were screened. Fifteen of these facilities underwent mass screening in response to a reported diagnosis of COVID-19 in a resident (Table: “Response group” Facilities 1–15), while 13 of the facilities did not have a known active case of COVID-19 at the time of testing and underwent mass screening as a preventive measure (“Preventive group” Facilities 16-28).

Overall, 28.6% (n=821) of residents and 9.4% (n=264) of staff screened were confirmed to have COVID-19. The prevalence of COVID-19 infection was significantly higher among residents and staff of LTCFs screened in response to a known case, compared to LTCFs screened for preventive purposes (Residents: Response group 47.2% vs. Preventive group 1.5%, p<0.0001; Staff: Response group 12.8% vs. Preventive group 1.7%, p<0.0001; **Table**). All 15 (100%) of the Response group facilities had a substantial proportion of both residents and staff with SARS-CoV-2 infection (Residents: median 46% [IQR 27% − 73%] infected; Staff: median 9.4% [IQR 5.9% − 20%]). Although the infection rate was significantly lower in the Preventive Group facilities, 7 (54%) of 13 facilities still had at least one resident who was SARS-CoV-2 infected and 4 (31%) had at least one infected staff member.

**Table 1.** COVID-19 Positivity, hospitalizations and deaths among Long-term Care Facility Staff and Residents screened during mass screening outreach in Fulton County, Georgia (March–May 2020)

| Facility (Date Screened) | Resident Count | Positive n (%)<sup>a</sup> | Hosp. n (%)<sup>b</sup> | Deaths n (%)<sup>b</sup> | Staff Count | Positive n (%)<sup>a</sup> | Hosp. n (%)<sup>b</sup> | Deaths n (%)<sup>b</sup> | Total screened | Positive n (%)<sup>a</sup> | Hosp. n (%)<sup>b</sup> | Deaths n (%)<sup>b</sup> |
| --- | --- | --- | --- | --- | --- | --- | --- | --- | --- | --- | --- | --- |
| **Response Group** |  |  |  |  |  |  |  |  |  |  |  |  |
| LTCF 1 (3/31/20) | 176 | 131 (74.4) | 18 (13.7) | 21 (16.0) | 74 | 40 (54.1) | 0 | 0 | 250 | 171 (68.4) | 18 (10.5) | 21 (12.3) |
| LTCF 2 (4/3/20) | 63 | 52 (82.5) | 17 (32.7) | 19 (36.5) | 81 | 32 (39.5) | 6 (18.8) | 0 | 144 | 84 (58.3) | 23 (27.4) | 19 (22.6) |
| LTCF 3 (4/5/20) | 69 | 17 (24.6) | 4 (23.5) | 2 (11.8) | 135 | 11 (8.1) | 0 | 0 | 204 | 28 (13.7) | 4 (14.3) | 2 (7.1) |
| LTCF 4 (4/8/20) | 67 | 49 (73.1) | 19 (38.8) | 12 (24.5) | 56 | 31 (55.4) | 2 (6.5) | 0 | 123 | 80 (65.0) | 21 (26.3) | 12 (15.0) |
| LTCF 5 (4/11/20) | 38 | 28 (73.7) | 9 (32.1) | 7 (25.0) | 61 | 13 (21.3) | 0 | 0 | 99 | 41 (41.4) | 9 (22.0) | 7 (17.1) |
| LTCF 6 (4/13/20) | 78 | 12 (15.4) | 2 (16.7) | 7 (58.3) | 199 | 13 (6.5) | 1 (7.7) | 0 | 277 | 25 (9.0) | 3 (12.0) | 7 (28.0) |
| LTCF 7 (4/15/20) | 112 | 52 (46.4) | 5 (9.6) | 1 (1.9) | 116 | 17 (14.7) | 0 | 0 | 228 | 69 (30.3) | 5 (7.2) | 1 (1.4) |
| LTCF 8 (4/16/20) | 88 | 40 (45.5) | 7 (17.5) | 3 (7.5) | 126 | 16 (12.7) | 2 (12.5) | 0 | 214 | 56 (26.2) | 9 (16.1) | 3 (5.4) |
| LTCF 9 (4/19/20) | 167 | 124 (74.3) | 32 (25.8) | 15 (12.1) | 130 | 25 (19.2) | 0 | 0 | 297 | 149 (50.2) | 32 (21.5) | 15 (10.1) |
| LTCF 10 (4/22/20) | 96 | 25 (26.0) | 5 (20.0) | 3 (12.0) | 104 | 4 (3.8) | 0 | 0 | 200 | 29 (14.5) | 5 (17.2) | 3 (10.3) |
| LTCF 11 (4/28/20) | 196 | 55 (28.1) | 7 (12.7) | 6 (10.9) | 150 | 4 (2.7) | 0 | 0 | 346 | 59 (17.1) | 7 (11.9) | 6 (10.2) |
| LTCF 12 (4/30/20) | 196 | 48 (24.5) | 6 (12.5) | 2 (4.2) | 252 | 3 (1.2) | 0 | 0 | 448 | 51 (11.4) | 6 (11.8) | 2 (3.9) |
| LTCF 13 (5/7/20) | 94 | 28 (29.8) | 8 (28.6) | 4 (14.3) | 75 | 4 (5.3) | 0 | 0 | 169 | 32 (18.9) | 8 (25.0) | 4 (12.5) |
| LTCF 14 (5/11/20) | 81 | 46 (56.8) | 6 (13.0) | 6 (13.0) | 106 | 10 (9.4) | 1 (10.0) | 0 | 187 | 56 (29.9) | 7 (12.5) | 6 (10.7) |
| LTCF 15 (5/14/20) | 184 | 97 (52.7) | 26 (26.8) | 23 (23.7) | 279 | 26 (9.3) | 3 (11.5) | 0 | 463 | 123 (26.6) | 29 (23.6) | 23 (18.7) |
| **Preventive Group** |  |  |  |  |  |  |  |  |  |  |  |  |
| LTCF 16 (4/2/20) | 287 | 1 (0.3) | 0 | 0 | 270 | 0 | 0 | 0 | 557 | 1 (0.2) | 0 | 0 |
| LTCF 17 (4/29/20) | 102 | 1 (1.0) | 0 | 0 | 0 | 0 | 0 | 0 | 102 | 1 (1.0) | 0 | 0 |
| LTCF 18 (5/5/20) | 26 | 4 (15.4) | 4 (100.0) | 3 (75.0) | 8 | 0 | 0 | 0 | 34 | 4 (11.8) | 4 (100.0) | 3 (75.0) |
| LTCF 19 (5/6/20) | 64 | 0 | 0 | 0 | 64 | 0 | 0 | 0 | 128 | 0 | 0 | 0 |
| LTCF 20 (5/11/20) | 73 | 0 | 0 | 0 | 46 | 2 (4.3) | 0 | 0 | 119 | 2 (1.7) | 0 | 0 |
| LTCF 21 (5/13/20) | 78 | 1 (1.3) | 1 (100.0) | 0 | 100 | 1 (1.0) | 0 | 0 | 178 | 2 (1.1) | 1 (50.0) | 0 |
| LTCF 22 (5/18/20) | 46 | 0 | 0 | 0 | 43 | 0 | 0 | 0 | 89 | 0 | 0 | 0 |
| LTCF 23 (5/27/20) | 35 | 0 | 0 | 0 | 19 | 0 | 0 | 0 | 54 | 0 | 0 | 0 |
| LTCF 24 (5/27/20) | 48 | 6 (12.5) | 0 | 0 | 76 | 10 (13.2) | 0 | 0 | 124 | 16 (12.9) | 0 | 0 |
| LTCF 25 (5/28/20) | 218 | 2 (0.9) | 0 | 0 | 100 | 2 (2.0) | 1 (50.0) | 1 (50.0) | 318 | 4 (1.3) | 1 (25.0) | 1 (25.0) |
| LTCF 26 (5/29/20) | 87 | 2 (2.3) | 0 | 0 | 97 | 0 | 0 | 0 | 184 | 2 (1.1) | 0 | 0 |
| LTCF 27 (5/29/20) | 1 | 0 | 0 | 0 | 30 | 0 | 0 | 0 | 31 | 0 | 0 | 0 |
| LTCF 28 (5/29/20) | 98 | 0 | 0 | 0 | 6 | 0 | 0 | 0 | 104 | 0 | 0 | 0 |
| **Total** | **2868** | **821 (28.6)** | **176 (21.4)** | **134 (16.3)** | **2803** | **264 (9.4)** | **16 (6.1)** | **1 (0.4)** | **5671** | **1085 (19.1)** | **192 (17.7)** | **135 (12.4)** |
| Total Response screening | 1705 | 804 (47.2) | 171 (21.3) | 131 (16.3) | 1944 | 249 (12.8) | 15 (6.0) | 0 | 3649 | 1053 (28.9) | 186 (17.7) | 131 (12.4) |
| Total Preventive screening | 1163 | 17 (1.5) | 5 (29.4) | 3 (17.6) | 859 | 15 (1.7) | 1 (6.7) | 1 (6.7) | 2022 | 32 (1.6) | 6 (18.8) | 4 (12.5) |
| p value<sup>c</sup> |  | **<0.0001** | 0.38 | 0.75 |  | **<0.0001** | 1.0 | 0.06 |  | **<0.0001** | 0.82 | 1.0 |

<sup>a</sup> Proportion among all persons screened
<sup>b</sup> Proportion among COVID-19 positive persons
<sup>c</sup> Fisher's exact test used. Significant p values bolded for ease of interpretation.

Of the 1,085 individuals (residents and staff) diagnosed with COVID-19, 18% (n=192) required hospitalization and 12% died (n=135). Although hospitalization rates did not differ significantly among COVID-positive residents in the Response group and those in the Preventive group (21% vs 29%, p=0.38), the magnitude of hospitalizations was overwhelmingly among residents and staff in LTCFs in the Response group (n=186 of 191 hospitalized, 97%). Similarly, among the overall 135 deaths among COVID-positive individuals across all 28 LTCFs, the majority (n=131, 97%) occurred among residents living in Response group facilities.

## Discussion

Long-term care facilities have borne a disproportionate burden of COVID-19 morbidity and mortality. In this study, we analyzed data collected from mass screenings conducted among 28 LTCFs in Fulton County, Georgia, both as a response to a confirmed case and as a preventive measure before active infections were identified. We found that more than 1 in 4 LTCF residents, and 1 in 10 LTCF staff, had SARS-CoV-2 infection. However, in facilities where testing was conducted only after a case had been identified, a dramatically greater proportion of residents (47% vs 1.5%) and staff (13% vs 1.7%) were found to be infected, suggesting SARS-CoV-2 infection was already widespread by the time a single case was diagnosed through passive means. Encouragingly, among LTCFs screened before a case was passively diagnosed, only about 2% of residents and 2% of staff were found to be positive, and few deaths were reported. These data support the role of active screening in LTCFs, in addition to routine infection prevention and control measures, as part of the COVID-19 response.

In our study, LTCFs with at least one COVID-19 case passively diagnosed had a median of 46% of its residents infected and 9.4% staff. This finding is consistent with data from other settings that suggest 10–100 cases may be present for every 1 case diagnosed [9]. An additional important finding from our study is that although facilities in the Preventive Group had lower SARS-CoV-2 infection rates, the majority still had at least one infected resident and one-third had at least one infected staff member.

These data support recent guidance provided by the Centers for Medicare and Medicaid Services (CMS), which recommends weekly testing for LTCF staff regardless of the presence of COVID-19 cases in the LTCF [10]. CMS further recommends that if a SARS-CoV-2 infection has been identified within the facility, weekly testing should be conducted of residents until all have tested negative.

The lower rate of infection in the Preventive group compared to the Response group demonstrates the rapid spread of COVID-19 that can occur if identification of infection is delayed until the onset of symptoms. Mass screening can diagnose residents and staff who are infected with SARS-CoV-2 but are in an asymptomatic or presymptomatic phase. Identification of such individuals can lead to the prevention of additional infections by isolating residents and excluding staff that are infected, initiating contact tracing, and increasing infection prevention measures including cleaning and sterilization procedures.

One concern with conducting serial RT-PCR testing of infected individuals until they test negative is the uncertainty of whether a person who tests positive is actively infected and infectious, or whether a positive test represents non-viable viral material. A recent study found that the SARS-CoV-2 virus was unable to be cultured in specimens that tested positive by RT-PCR 8 days after symptom onset [11], though individuals diagnosed with COVID-19 can continue to have detectable virus by PCR for as long as 6 weeks [12]. Another challenge to COVID-19 surveillance in some LTCFs is the fear of negative public perceptions when reporting infections. In response to this issue and COVID-19 outbreaks in LTCFs in Fulton County, the FCBOH has worked to improve collaboration and communication between LTCFs and other governmental entities while providing support through stocking of personal protective equipment (PPE), training by state or locally designated infection prevention specialists, and facilitating opportunities for LTCFs to meet and discuss their strategies to prevent additional infections.

One limitation of this study is that LTCFs were screened based on reports of COVID-19 infections or requests by the facilities and were not selected at random to provide a representative sample. Nonetheless, these facilities represent 48% of licensed LTCFs and 44% of the total bed capacity in Fulton County [13]. Due to low testing capacity in the early phase of the COVID response, most LTCFs screened early in the study period (April) were those with a known case (Response Group). Preemptive screening began in May when testing capacity increased with the addition of the National Guard to the COVID-19 screening efforts.

While typical case investigations for COVID-19 require contacting individuals directly, we were unable to contact COVID-19 positive residents of LTCFs to obtain epidemiologic details such as the date of onset of symptoms or potential close contacts; we relied heavily on LTCFs to assist us in completing individual case investigations. Census lists provided by LTCFs and case reports from hospitals and medical examiners assisted in the identification and retroactive linkage of cases in residents and staff that were not reported by LTCFs and their subsequent inclusion in this analysis. Lastly, there may be several unmeasured factors that could account for differences between the LTCF groups, such as resident to staff ratio, resident and staff adherence to IPC measures, prevalence of comorbid conditions especially among residents, facility airflow design and maintenance practices, and degree of functional impairment that may predispose to more frequent and closer resident-staff interaction.

The COVID-19 pandemic has underscored the fact that residents and staff of LTCFs represent a highly vulnerable population. Critical analyses of the mass screening efforts conduced in Fulton County provide support for active screening to identify cases early and increase infection control and prevention measures. Together, these measures can reduce spread of SARS-CoV-2 and can provide residents and staff with the best possible chance for a successful outcome from this devastating illness.

## Data Availability

Data was gathered by the Fulton County Board of Health and can be obtained through data use agreements and approval by the data steward at the Fulton County Board of Health.

## Acknowledgements

We are grateful to the Fulton County Board of Health staff for their tireless efforts in responding to the COVID-19 pandemic, including testing and case management. We thank the facilities who requested testing and all residents and staff who participated.

## Disclaimers

The findings and conclusions in this manuscript are those of the authors and do not necessarily represent the official position of the funders. The funders had no role in study design, data collection and analysis, decision to publish, or preparation of the manuscript.

